# Machine Learning Versus Logistic Regression for Propensity Score Estimation: A Benchmark Trial Emulation Against the PARADIGM-HF Randomized Trial

**DOI:** 10.1101/2025.06.16.25329708

**Authors:** Kaicheng Wang, Lindsey Rosman, Haidong Lu

## Abstract

Machine learning (ML) algorithms are increasingly used to estimate propensity score with expectation of improving causal inference. However, the validity of ML-based approaches for confounder selection and adjustment remains unclear. In this study, we emulated the device-stratified secondary analysis of the PARADIGM-HF trial among U.S. veterans with heart failure and implanted cardiac devices from 2016 to 2020. We benchmarked observational estimates from three propensity score approaches against the trial results: (1) logistic regression with pre-specified confounders, (2) generalized boosted models (GBM) using the same pre-specified confounders, and (3) GBM with expanded covariates and automated feature selection. Logistic regression-based propensity score approach yielded estimates closest to the trial (HR = 0.93, 95% CI 0.61 - 1.42; 23-month RR = 0.86, 95% CI 0.57 - 1.24 vs. trial HR = 0.81, 95% CI 0.61-1.06). Despite better predictive performance, GBM with pre-specified confounders showed no improvement over the logistic regression approach (HR = 0.97, 95% CI 0.68 – 1.37; RR = 0.96, 95% CI 0.89 – 1.98). Notably, GBM with expanded covariates and data-driven automated feature selection substantially increased bias (HR = 0.61, 95% CI 0.30 - 1.23; RR = 0.69, 95% CI 0.36 - 1.04). Our findings suggest that ML-based propensity score methods do not inherently improve causal estimation—possibly due to residual confounding from omitted or partially adjusted variables—and may introduce overadjustment bias when combined with automated feature selection, underscoring the importance of careful confounder specification and causal reasoning over algorithmic complexity in causal inference.

## INTRODUCTION

Observational studies rely on appropriate confounding adjustment to estimate causal effects, yet confounder selection practices remain suboptimal. A review of high-impact medical and epidemiologic journals found that, in 2023, 41.5% of observational studies failed to justify their confounder set, while 16.5% relied solely on data-driven criteria such as change-in-estimate or stepwise regression.(1) Such practices are concerning because valid causal inference requires subject-matter knowledge to identify confounders—common causes of both exposure and outcome—thereby approximating conditional exchangeability.(2)

Machine learning (ML) algorithms, such as generalized boosted models (GBM) and high-dimensional propensity score, are increasingly used for propensity score estimation, and sometimes to select covariates directly from electronic health record (EHR) data.(3-7) These algorithms offer several advantages, including automated feature selection and the ability to model non-linear relationships and complex interactions without parametric assumptions.(8) This can improve discrimination and calibration in predictive performance, leading some investigators to expect similar improvements from causal inference applications.(9) However, optimizing predictive accuracy does not guarantee bias reduction in causal inference.(10, 11) Automated feature selection, without any pre-specifications based on subject-matter knowledge, can introduce overadjustment bias(12) by inadvertently including mediators, colliders or instrumental variables. Although prior studies(8, 13, 14) have evaluated the performance of ML-based propensity scores, most have relied on simulations with well-defined data-generating mechanisms and simplified causal structures that may not reflect real-world data-generating processes.

In this study, we applied benchmarking methods—comparing results of observational analyses with randomized controlled trial that addressing identical research questions—to provide a more rigorous evaluation of analytic choices in realistic settings.(15-17) Specifically, we emulated the device-stratified secondary analysis(18) of the PARADIGM-HF trial(19) which compared sacubitril/valsartan with enalapril among patients with heart failure (HF), restricted to those with an implantable cardioverter defibrillator (ICD) or cardiac resynchronization therapy defibrillator (CRT-D). Importantly, we benchmarked observational estimates against the index PARADIGM-HF trial results using three propensity score approaches: (1) logistic regression with pre-specified confounders, (2) generalized boosted model (GBM) with the same pre-specified confounder set, and (3) GBM with additional predictors via automated feature selection. Our objective was to assess whether ML-based propensity score estimation together with automated feature selection improves or worsens alignment with results from randomized trial compared with traditional regression-based estimation.

## METHODS

### Benchmark Trial

The PARADIGM-HF(19) was a multicenter, randomized, double-blind trial comparing the efficacy of sacubitril/valsartan with enalapril among 8,442 patients with chronic HF (NCT01035255). All-cause mortality was a prespecified endpoint, along with cardiovascular-specific mortality and HF hospitalization. For benchmarking, we used a secondary analysis(18) restricted to 1,243 participants who had an ICD or CRT-D at baseline, the same device-defined HF population we emulated to account for possible effect modification by device implantation. In this subgroup, sacubitril/valsartan was associated with a 19% lower risk of all-cause mortality compared to enalapril (HR = 0.81; 95% CI 0.61 - 1.06).

### Target Trial Specification and Observational Emulation

#### Study Design

We specified a hypothetical trial between January 1, 2016 and September 30, 2020 with eligibility criteria, treatment strategies, and the outcome that mirrored the device-stratified secondary analysis of PARADIGM-HF, using data from the VA Corporate Data Warehouse (CDW).(20) Key target trial protocol components are summarized in **Supplemental Table 1**.

#### Eligibility Criteria

We included veterans aged 18 years and older with an HF diagnosis (International Classification of Diseases, Ninth or Tenth Revision codes), left ventricular ejection fraction (LVEF) ≤ 35%, and ICD or CRT-D implantation between January 1, 2010 and September 30, 2020. Prior to the index date, patients were required to have a B-type natriuretic peptide (BNP) ≥ 150 pg/mL or N-terminal pro B-type natriuretic peptide (NT-proBNP)≥ 600 pg/mL, and to have received at least one guideline-directed medical therapy (β-blocks, mineralocorticoid receptor antagonists [MRAs) and sodium-glucose transport protein 2 inhibitors [SGLT2i]). Patients with a left ventricular assist device or a history of heart transplant were excluded.

#### Treatment Strategies

Sacubitril/valsartan prescriptions were obtained from the CDW outpatient pharmacy domain. To reflect real-world clinical practice, rather than enalapril alone, the comparator group included all ACEIs (e.g., -pril) and ARBs (e.g., - sartan, including valsartan), as these agents share pharmacological mechanisms and are guideline-recommended for HF treatment. The baseline index date (i.e., time zero) was defined as the first prescription after meeting all eligibility criteria for sacubitril/valsartan initiators, and the first eligible prescription (re)fill during the study period for ACEI/ARB users.

The 2016 AHA/ACC HF guideline(21) recommended augmenting ACEI/ARB therapy with a neprilysin inhibitor (sacubitril) to further reduce morbidity and mortality, with *de novo* initiation of sacubitril/valsartan recommended beginning in 2022.(22) Accordingly, most sacubitril/valsartan initiators in this study were prevalent users(23) of an ACEI/ARB prior to the index date. Hence, in the current observational analyses, the treatment strategies include those who augment an ACEI/ARB with sacubitril versus those who continue ACEI/ARB therapy. This treatment decision design(24) is consistent with the two-week enalapril run-in period in PARADIGM-HF. Prevalent users in this analysis were defined as patients who had filled an ACEI/ARB prescription within 180 days prior to either augmenting to sacubitril/valsartan or refilling an ACEI/ARB.(25) Adherence was defined as receiving a refill within 30 days after the end of supply.

#### Outcome and Follow-up

The primary outcome was all-cause mortality, identified from the VA Death ascertainment file that passively surveilled through VA Master Person Index(26), administrative data, and Social Security Administration, with a 98.3% sensitivity and 97.6% exact agreement with National Death Index.(27) Therefore, we assumed no misclassification of the outcome and no loss to follow-up. Patients were followed from the baseline until the date of death, 51 months after baseline, or administrative end of the study by September 30, 2020, whichever happened first.

#### Potential Confounders Based on Subject-Matter Knowledge

Potential confounders (**Supplemental Table 2**) were identified based on subject-matter knowledge and previous literature.(28) We assumed treatment strategies were exchangeable conditional on baseline confounders, including trial sequence number, demographics (age, gender, race, ethnicity), social determinants of health (cigarette use, area deprivation index [ADI]), healthcare-seeking behavior (flu vaccination or HF hospitalization in the past 12 months), clinical characteristics (body mass index [BMI], systolic blood pressure [SBP], LVEF, estimated glomerular filtration rate [eGFR], serum/plasma potassium, diagnosis of atrial fibrillation, cardiomyopathy, chronic kidney disease [CKD], chronic obstructive pulmonary disease [COPD], diabetes, hypertension, depression, history of myocardial infraction [MI], peripheral artery disease [PAD] and valvular heart disease [VHD]), prescriptions (β-blockers, MRAs, SGLT2i, diuretics and digoxin), CRT, user type (new vs. prevalent user) and time since HF diagnosis.

#### Non-Confounding Predictors

Although high-dimensional data-driven approaches can incorporate hundreds of variables, fourteen additional predictors were included to illustrate the potential for causal model misspecification and bias when variables not meeting the definition of confounders are incorporated. These data-driven variables (*C*^*^) included strong predictors of the treatment assignment (residence [urban vs. rural], employment status, time since device implantation) and predictors of mortality not directly associated with treatment assignment (anemia, cerebral vascular conditions, deep vein thrombosis and pulmonary embolism [DVT/PE], dementia, dyslipidemia, influenza, non-pathological fracture, malignancy, obstructive sleep apnea [OSA], hemoglobin, and albumin).

#### Statistical Analyses

Baseline characteristics were summarized as frequencies and percentages, means and standardized deviations (SDs), or medians and interquartile ranges (IQRs) for non-normally distributed variables. For computational feasibility, continuous variables with missing values (ADI, hemoglobin, potassium, albumin) were imputed with medians, and categorical variables were coded as “Unknown”. Data were managed using SQL Server Management Studio 19.0, and statistical analyses were conducted using R version 4.4.1.

First, we estimated propensity scores using logistic regression on the pre-specified confounders. We performed an intention-to-treat analysis using stabilized inverse probability of treatment weights (IPTW). Covariate balance was assessed using standardized mean difference (SMD), with a value less than 0.1 indicating good balance between treatment groups. Weighted pooled logistic regression models were fit with flexible time-varying intercepts and treatment-by-time interaction to estimate 23-month (median follow-up time) cumulative risks. Risk differences (RDs) and risk ratios (RRs) were computed, with confidence intervals (CIs) obtained nonparametrically using 500 bootstrapping samples. Hazard ratios (HRs) were estimated using pooled logistic regression models without the interaction term, and the corresponding CIs were estimated through the robust variance estimator.

Next, propensity scores were estimated using generalized boosted model (GBM)(29, 30), a commonly used machine learning algorithm, with the same pre-specified confounders. The model began with a shrinkage parameter of 0.1 and 1,000 iterations. Hyperparameters, including shrinkage (0.3, 0.1, 0.05, 0.01 and 0.005), interaction.depth (2, 3, 5 and 7) and n.minobsinnode (10, 20, 30, 50, 75, 100 and 150) were tuned based on 10-fold cross-validation errors.

Finally, to illustrate the potential risk of data-drive automated confounder selection, we repeated the GBM analysis after adding 14 predictors to the pre-specified confounder set. Although treatment classification metrics such as the area under the receiver operating characteristic curve (AUC) are not relevant for causal inference, and can be misleading when interpreted as measures of confounding control, we reported AUCs for all three propensity score approaches for demonstrative purposes only.

We then informally benchmarked the intention-to-treat estimates from the target trial emulation, using each propensity score method, against the intention-to-treat all-cause mortality result from the PARADIGM-HF device subgroup.(18) We assessed whether the GBM-based estimates together with data-driven automated confounder selection aligned more closely with, or diverged further from the trial findings compared with logistic regression-based estimates.

## RESULTS

A total of 583 sacubitril/valsartan initiators and 1,864 ACEI/ARB users between January 1, 2016 and September 30, 2020 were analyzed (**Table 1 & Fig. 1**). The median follow-up time was 23.0 [12.0 – 38.0] months. Compared to those receiving ACEI/ARB, the sacubitril/valsartan group was younger (mean [SD], 68.6 [8.6] vs. 70.2 [8.2] years), had a lower proportion of White individuals (68.3% vs. 74.0%). The sacubitril/valsartan group had a higher proportion of patients diagnosed with cardiomyopathy (84.4% vs. 78.5%), depression (40.5% vs. 36.4%), and prescribed MRAs (48.9% vs. 35.6%), diuretics (68.4% vs. 60.4%) and digoxin (11.5% vs. 9.0%), and a lower proportion of patients with CRT-D (44.6% vs. 49.6%), any HF hospitalizations (44.6% vs. 54.8%) or influenza vaccine (71.7% vs. 75.3%) in the past 12 months, compared to the ACEI/ARB group. Additionally, patients receiving sacubitril/valsartan had a longer duration since the initial diagnose of HF (median [IQR], 80.0 [38.5 – 147.5] vs. 62.0 [14.0-125.0] months) and higher proportion of prevalent users (93.1% vs. 83.6%).

**Figure 1.**
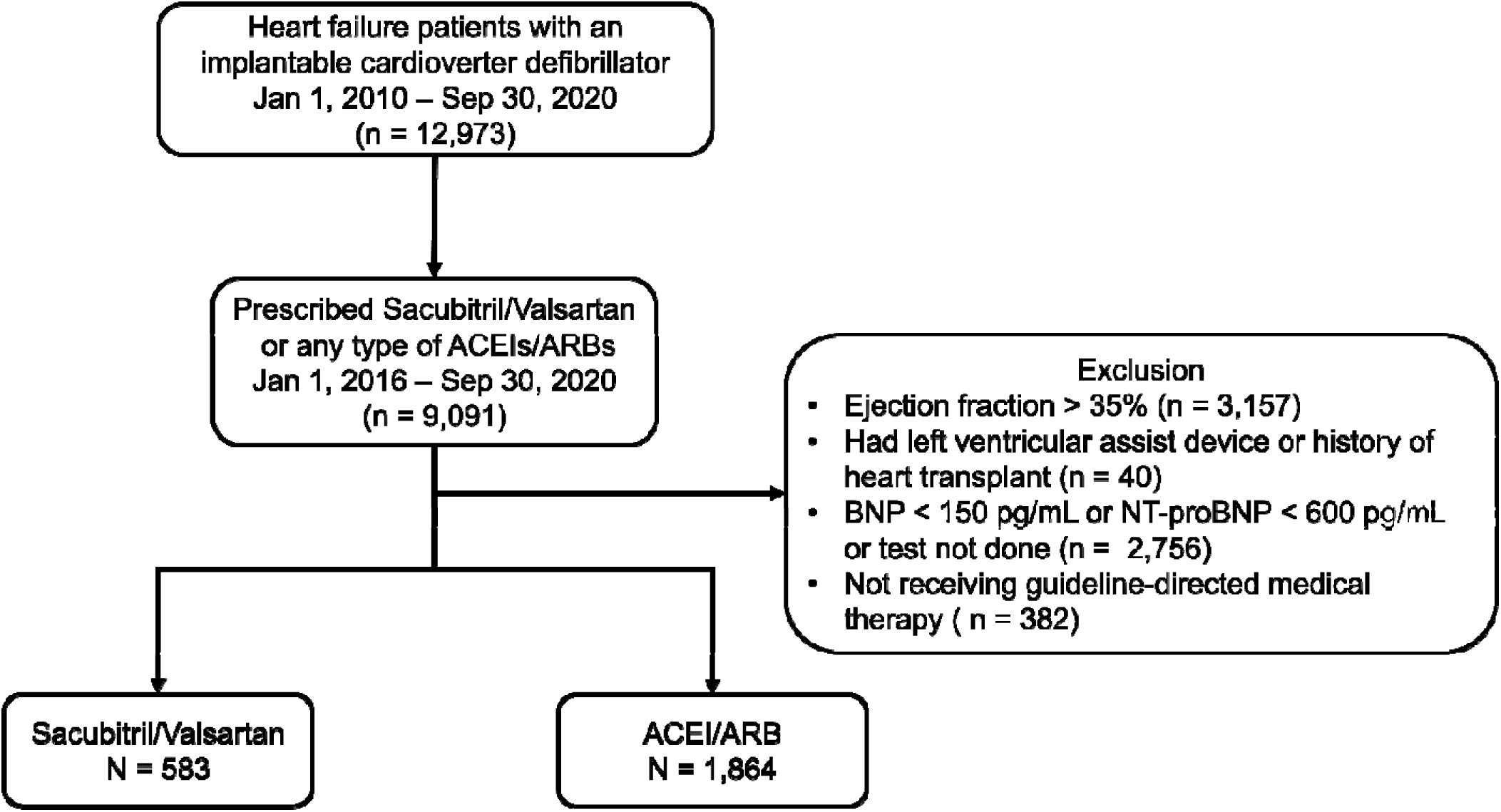
Study Flow Chart. Abbreviations: ACEIs/ARBs, angiotensin converting enzyme inhibitors and angiotensin II receptor blockers; BNP, brain natriuretic peptide; NT-proBNP, N-terminal pro-brain natriuretic peptide.

**Table 1.**
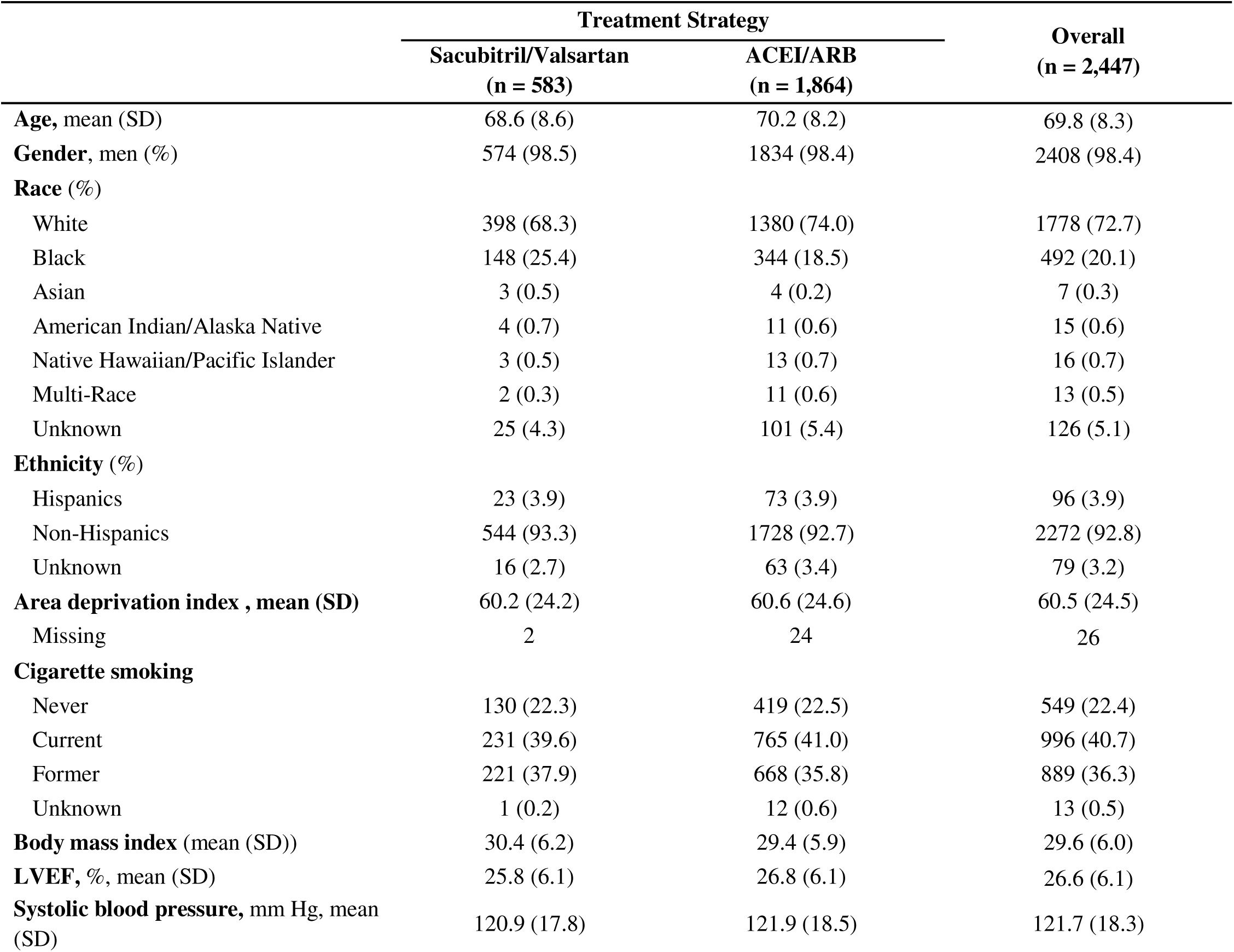
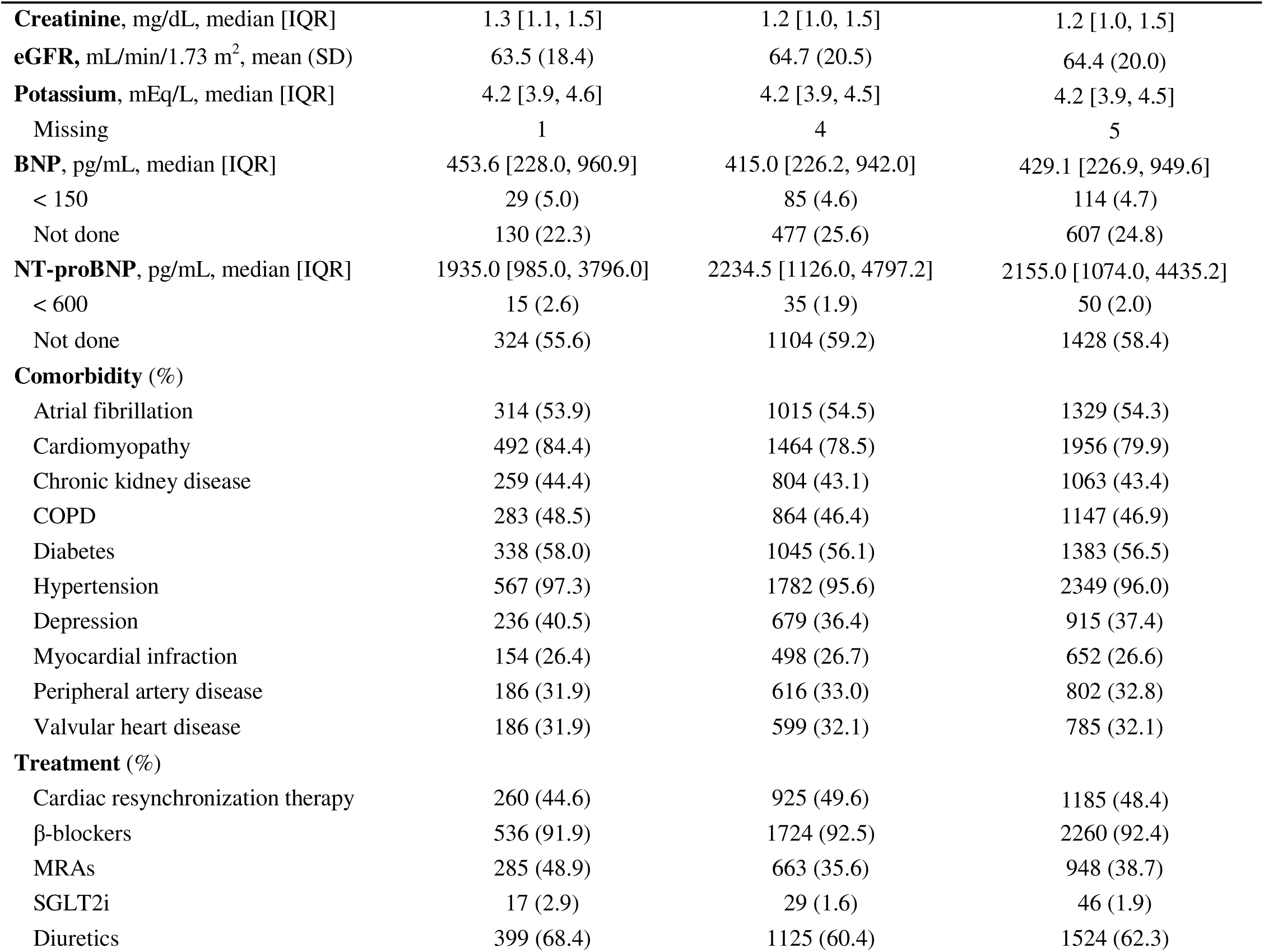
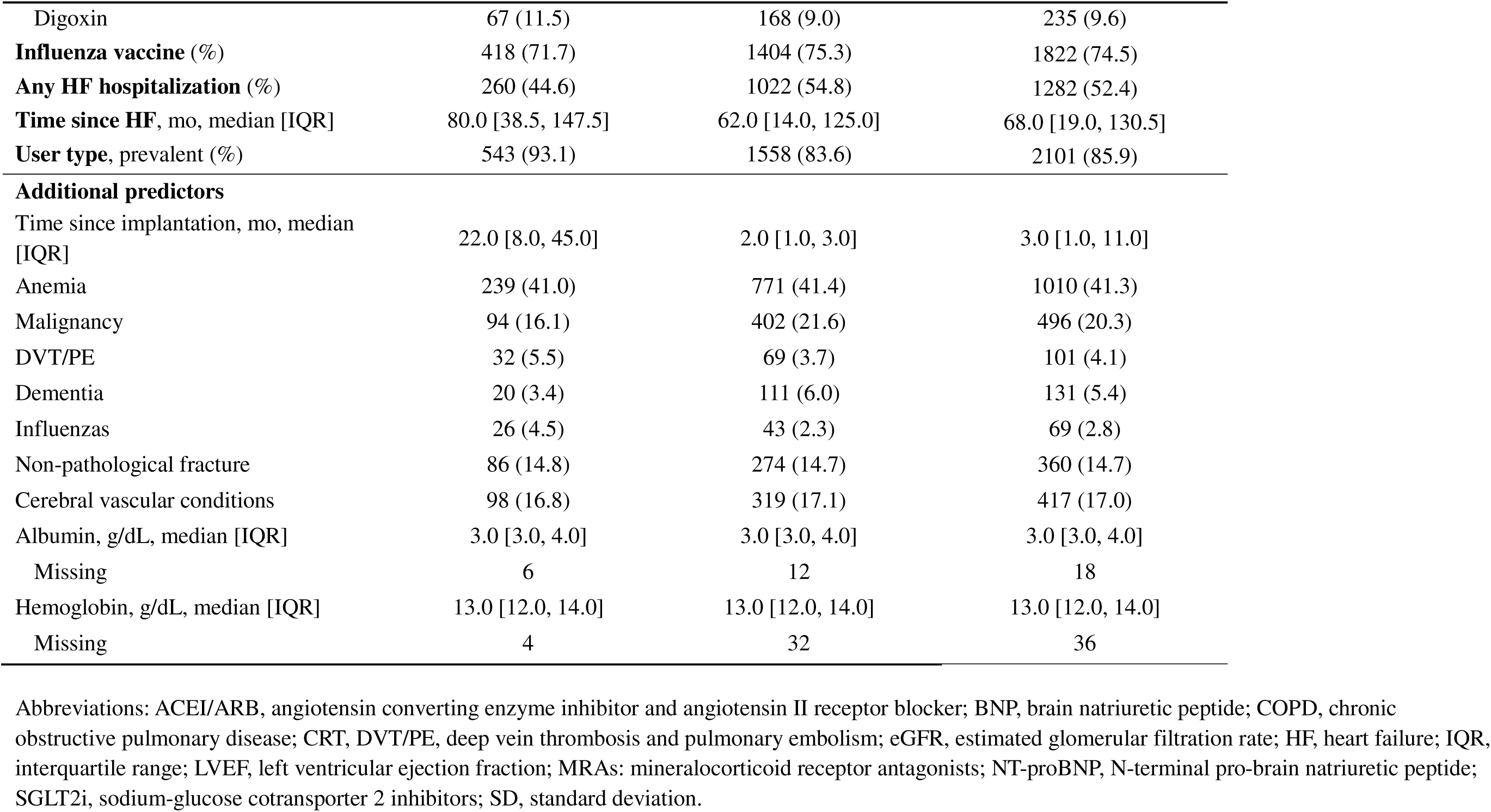
Baseline Characteristics of Heart Failure Patients with Implantable Cardioverter Defibrillators Prescribed Sacubitril/Valsartan or ACEI/ARB in the U.S. Department of Veterans Affairs, 2016 – 2020.

### Estimates From Pre-Specified Confounders and Logistic Regression-Based Propensity Score

The distribution of propensity scores is visualized in **Fig, 1a**, with an AUC 0.79 (95% CI, 0.77 - 0.81) for treatment classification. After applying the IPTW, baseline covariates were well balanced (**Supplemental Fig. 1**). The 23- month cumulative incidence of all-cause mortality (**Fig. 1b**) was 7.7% (95% CI, 5.0 – 10.8%) in the sacubitril/valsartan group compared to 9.0% (95% CI, 7.5 – 10.4%) in the ACEI/ARB group, leading to an RD of - 1.3% (95% CI, -4.1 – 2.0%), RR 0.86 (95% CI, 0.57 – 1.24), and HR 0.93 (95% CI, 0.61 – 1.42) during the entire follow-up period (**Table 2**).

**Table 2.** Intention-to-Treat Estimates of Cumulative Incidence, Risk Difference, Risk Ratio and Hazard Ratio for All-Cause Mortality from an Emulated Target Trial of Sacubitril/Valsartan versus ACEI/ARB, Benchmarked Against the Device-Restricted Secondary Analysis of the Index PARADIGM-HF Trial.

|  | 23-months cumulative incidence |  |  |  | HR | AUC (95% CI) |
| --- | --- | --- | --- | --- | --- | --- |
|  | Intervention, %<br>(95% CI) | Comparator, %<br>(95% CI) | RD<br>(95% CI) | RR<br>(95% CI) | (95% CI) |  |
| Index trial |  |  |  |  | 0.81 (0.61, 1.06) |  |
| Emulated target trial |  |  |  |  |  |  |
| Logistic regression, pre-specified confounders | 7.7 (5.0, 10.8) | 9.0 (7.5, 10.4) | -1.3 (-4.1, 2.0) | 0.86 (0.57, 1.24) | 0.93 (0.61, 1.42) | 0.79 (0.77, 0.81) |
| GBM, pre-specified confounders | 8.6 (8.0, 16.1) | 8.9 (7.1, 10.2) | -0.3 (-1.0, 7.8) | 0.96 (0.89, 1.98) | 0.97 (0.68, 1.37) | 0.81 (0.79, 0.83) |
| GBM, pre-specified confounders* | 6.9 (5.6, 15.0) | 7.0 (4.4, 9.5) | -0.3 (-0.7, 7.2) | 0.96 (0.91, 2.10) | 0.97 (0.68, 1.38) |  |
| GBM, additional predictors | 6.2 (3.3, 10.5) | 9.0 (7.6, 10.5) | -2.8 (-6.1, 1.5) | 0.69 (0.36, 1.18) | 0.61 (0.30, 1.23) | 0.95 (0.94, 0.97) |

Abbreviations: ACEI/ARB, angiotensin converting enzyme inhibitors and angiotensin II receptor blockers; CI, confidence interval; GBM, generalized boosted model; HR, hazard ratio; logistic regression; RD, risk difference; RR, risk ratio.

### Estimates From Pre-Specified Confounders and GBM-Based Propensity Score

The distribution of GBM-based propensity scores is visualized in **Fig. 1c**, with an AUC of 0.81 (95% CI, 0.79 - 0.83). After applying the weights, the SMDs were greater than 0.1 for cardiomyopathy (0.113), diuretics (0.112) and user type (0.129). Variable importance is visualized in **Supplemental Fig. 2a**. Gender, ethnicity, baseline hypertension and prescription of β-blockers and SGLT2i were not used in any boosted trees. The 23-month cumulative incidence of all-cause mortality (**Fig. 1d**) was 8.6% (95% CI, 8.0 – 16.1%) in the sacubitril/valsartan group vs. 8.9% (95% CI, 7.1 – 10.2%) in the ACEI/ARB group, corresponding to an RD of -0.3% (95% CI, -1.0 – 7.8%), RR 0.96 (95% CI, 0.89 – 1.98), and HR 0.97 (95% CI, 0.68-1.37). After additionally adjusted for cardiomyopathy, diuretic prescriptions and user type in the weighted pooled logistic regression model, the RR (0.96; 95% CI, 0.91 – 2.10) and HR (0.97; 95% CI 0.68 - 1.38) did not change substantially.

### Estimates From GBM-Based Propensity Score with Additional Predictors

After including 14 predictors in the GBM with automated feature selection, the histogram of propensity scores demonstrated poor overlap (**Fig. 1e**). The median [IQR] predicted probability of receiving sacubitril/valsartan was 0.934 [0.629 - 0.996] in the sacubitril/valsartan group and 0.042 [0.022 – 0.087] in the ACEI/ARB group. The AUC was 0.95 (95% CI, 0.94 -0.97). Among these additional predictors, time since device implant, employment status, residency, hemoglobin and albumin levels, and diagnosis of anemia, malignancy, cerebral vascular conditions, OSA and non-pathological fractures were deemed predictive of treatment assignment by the algorithm (**Supplemental Fig. 2b**). The 23-month RR (**Fig. 1f**) was 0.61 (95% CI, 0.30 - 1.23) with an overall HR of 0.63 (95% CI, 0.31 - 1.30).

### Informal Benchmarking

Compared to PARADIGM-HF participants, our sample was older (mean [SD]: 69.8 [8.3] vs. 64.6 [9.9]), had fewer females (1.6% vs. 13.5%), more Black patients (20.1% vs. 6.8%), lower baseline LVEF (26.6 [6.1] vs. 27.3 [6.3]), higher prevalence of hypertension (97.3% vs. 67.5%), and lower use of guideline-directed medical therapy such as β-blockers (92.4% vs. 96.6%) and MRAs (38.7% vs. 56.5%).

The point estimation and CI from the logistic regression-based propensity score analysis (HR = 0.93, 95% CI 0.61 - 1.42; RR = 0.86, 95% CI 0.57 - 1.24) aligned most closely with the trial result (HR = 0.81; 95% CI 0.61 - 1.06), while point estimates from a GBM-based model using the same set of *pre-specified* confounders were closer to the null (HR = 0.97, 95% CI 0.68 - 1.37; RR = 0.96, 95% CI 0.89 - 1.98). When using a data-driven GBM-based approach with automatic selection, which incorporated variables predictive of treatment assignment (e.g., employment status, time since implantation) or empirically associated with mortality (e.g., albumin, hemoglobin), and the estimate was biased toward the alternative (HR = 0.61, 95% CI 0.30 - 1.23; RR = 0.69, 95% CI 0.36 - 1.18).

## DISCUSSION

In this observational study, we benchmarked different propensity score estimation strategies in an observational setting, using sacubitril/valsartan versus ACEI/ARB therapy in HF patients with ICDs/CRT-Ds as an empirical case study. We applied three propensity score approaches: (1) logistic regression with pre-specified confounders, (2) GBM using the same pre-specified confounders, and (3) GBM incorporating additional predictors via automated feature selection. Estimates from logistic regression-based propensity scores aligned most closely with the trial result, whereas GBM-based approaches failed to improve the estimates and, when relying on automated feature selection, introduced bias.

Findings from this investigation highlight a key distinction between prediction and causal inference. ML algorithms aim to maximize predictive accuracy by discriminating between interventions and comparators. In our analysis, the GBM with additional predictors almost perfectly predicted treatment assignment (AUC = 0.95; 95% CI 0.94 – 0.97), but the propensity score distributions across groups were barely overlapped (**Fig. 2e**). Such separation may violate the positivity assumption(31-33), which requires that all patients have a non-zero probability to receive either treatment at every level of confounders. This data-driven approach led to biased effect estimates (HR = 0.61, 95% CI 0.30 - 1.23)–even when the predictive performance appears excellent. Thus, while ML algorithms such as GBM can help mitigate concerns about statistical model misspecification (i.e., incorrect functional forms and relationship between exposure, covariates and outcome),(34-36) they do not automatically address causal model misspecification (i.e., identify a sufficient set of confounders and avoid different types of overadjustment biases), as causal model specification requires subject-matter knowledge.

**Figure 2.**
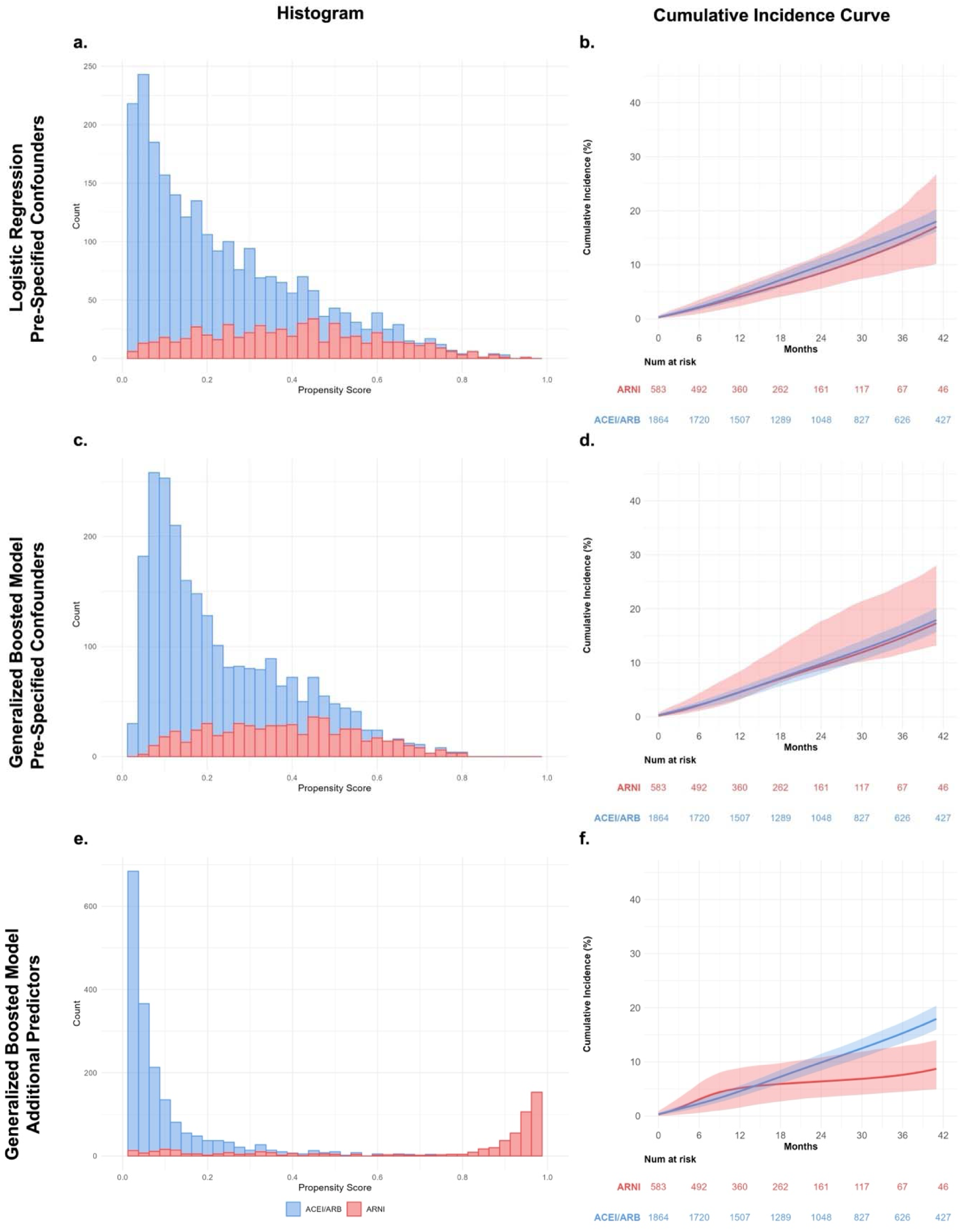
Propensity Score Distributions and Intention-to-Treat Cumulative Incidence Curves by Analytical Approach. Abbreviations: ACEI/ARB, angiotensin converting enzyme inhibitors and angiotensin II receptor blockers; ARNI, angiotensin receptor/neprilysin inhibitor.

Given these distinctions, claims about the advantages of ML for propensity score estimation require careful scrutiny. First, flexibility in modeling non-linearities and functional forms is not unique to ML algorithms: traditional regression can incorporate natural splines or restricted cubic splines(37), achieving similar improvements without the opacity of ML algorithms. Much of the enthusiasm for ML instead focuses on its ability to automate variable section–a feature that, for causal inference, can be problematic in both directions: to select and not to select.

On one side, automated selection may add inappropriate variables, such as colliders or instrumental variables. For instance, two overadjustment biases were identified in the GBM model with additional predictors through automated feature selection (**Fig. 3**): (a) overadjustment for a collider (albumin) and (b) overadjustment for a near-instrumental variable (time since implantation).

**Figure 3.**
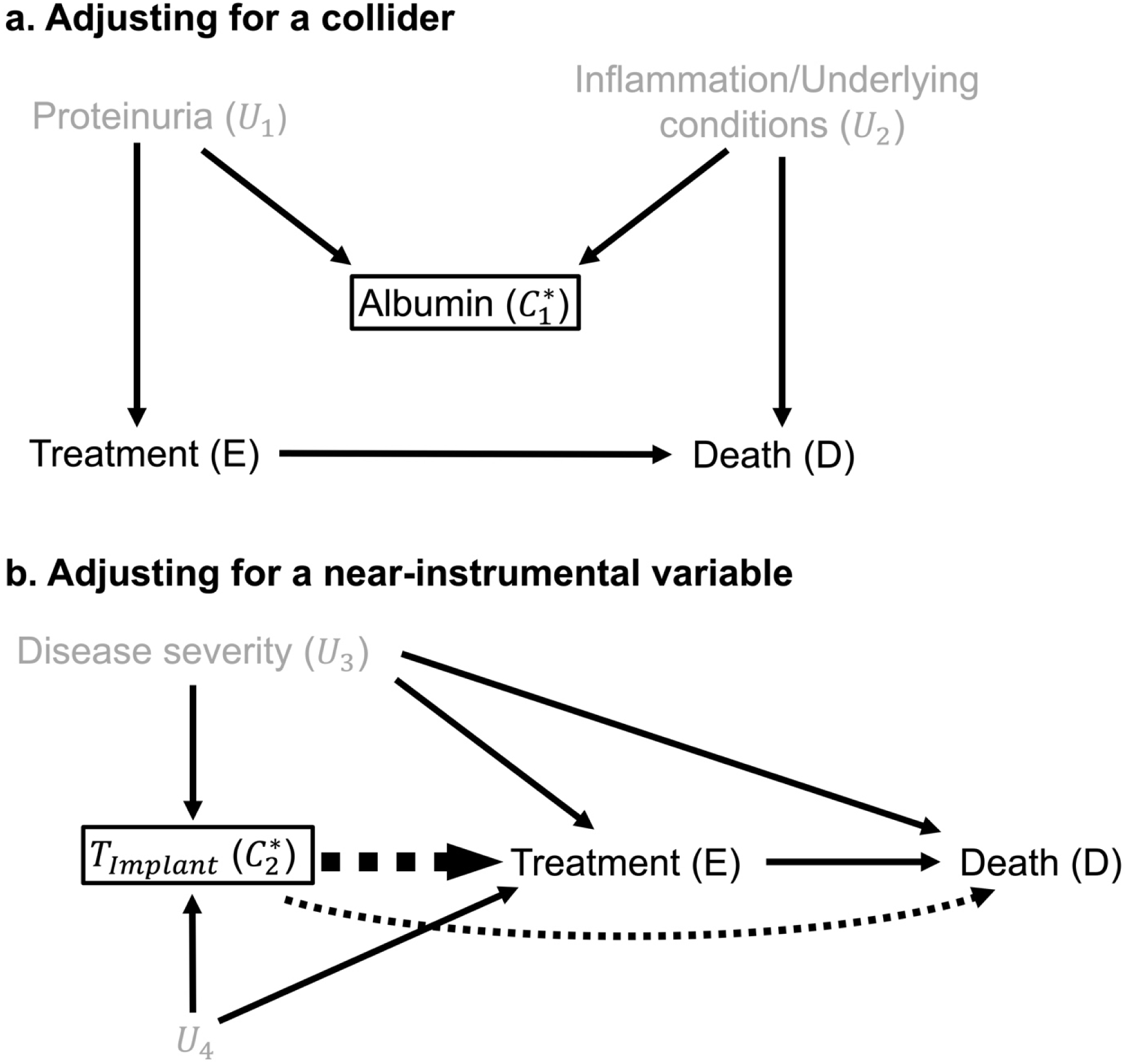
Directed Acyclic Graphs Illustrating Overadjustment Biases in the Data-Drive GBM-Based Propensity Score Approach. Black boxes indicate variables included in the propensity score model. Gray text represents unmeasured or unknown confounders. Dashed arrows denote non-causal paths implied by the data-driven GBM-based approach with arrow line thickness indicate strength of the association. Variables labeled *C*_*i*_^*^ represent GBM-imposed confounders.

In the first example, proteinuria *U*_1_ is the common cause of both hypoalbuminemia and initiation of ACEI/ARB therapy (*E*) to reduce the amount of protein that leaks into the urine. Meanwhile, systemic inflammation and other disease processes (unmeasured/unknown confounder, *U*_2_) also affect albumin levels and are independently associated with mortality *D*.(38) Adjusting for the collider albumin *C*_1_^*^ therefore opens a M bias pathway (*E* ← *U*_1_ → *C*_1_^*^ ← *U*_2_ → *D*) between treatment *E* and death *D*.

In the second example, time since implantation *C*_2_^*^ is an imperfect proxy for unmeasured HF severity *U*_3_, as ICD or CRT-D devices are reserved for HF patients who did not respond to guideline-directed pharmacotherapy. Even though time since implantation has no direct effect on treatment decision *E*, the algorithm included it as the strongest predictor of sacubitril/valsartan and imposed a causal path *C*_2_^*^ → *E*. As a shortcut feature(39, 40) in ML literature, adjusting for time since implantation could bias the estimation, if an unmeasured variable *U*_4_ exists as a common cause of both *C*_2_^*^ and *E* (overadjustment for collider). Moreover, because *C*_2_^*^ strongly predicts treatment, it may act as a near-instrumental variable(41, 42), where overadjustment bias (with a near-instrumental variable) may outweigh the limited precision gain from its weak association with the outcome. These forms of overadjustment biases likely explain why some prior studies, often simulation-based or secondary analysis of randomized trials, reported better performance with GBM and deep learning. Many of those studies either restricted analyses to pre-specified confounders (43-45) or did not explicitly consider scenarios where overadjustment could occur.

But does this mean that feeding ML algorithms only a carefully chosen set of confounders guarantees valid causal estimates? Not quite. Automated selection may also exclude important confounders if they weakly predict treatment. In our study, even when we supplied the same set of pre-specified confounders, the logistic regression-based approach (HR = 0.93, 95% CI 0.61 - 1.42; RR = 0.86, 95% CI 0.57 - 1.24) aligned more closely with the trial benchmark than the GBM-based approach (HR = 0.97, 95% CI 0.68 - 1.37; RR = 0.96, 95% CI 0.89 - 1.98). The importance plot (**Supplemental Fig. 2a**) showed β-blockers and SGLT2i—both Class 1A(22) therapies known to reduce HF hospitalizations and mortality—were dropped entirely from the GBM-based propensity model. Similarly, variables appearing only sporadically across trees, such as CKD with relatively low variable importance, may have been insufficiently adjusted.

That said, there remains at least one plausible advantage of ML in causal inference: detecting high-order interactions. Partial dependent plots, for example, can help visualize complex interactions between covariates and treatment assignment.(46) Yet, this benefit comes with caveats. Even if interactions are better captured, confounder selection must still be grounded in subject-matter knowledge, explicit causal reasoning, transparent statistical approach for confounder adjustment, rigorous diagnostics (e.g., propensity score distribution, covariate balance assessment, and variable importance if applicable), complemented by robustness assessments such as probabilistic bias analysis and positive/negative control outcomes.(47) Doubly-robust ML estimators through cross-fitting can also provide protection against model misspecification.(10)

This study has several limitations. First, we only conducted informal benchmarking without transportability analyses(48) from the trial population to the VA study sample. While the PARADIGM-HF trial reported a 19% reduction in all-cause mortality with sacubitril/valsartan vs. an ACEI (enalapril) among HF patients with devices, this efficacy may not be directly transportable due to heterogeneity of covariates distribution that may modify the treatment effect. However, without access to individual-level trial data, we could not formally transport the trial result to our study sample. Additionally, not all variables were available and perfectly measured. For instance, the New York Heart Association (NYHA) Functional Classifications, a key eligibility criterion in the trial, was not captured in our EHR data. Neither could we differentiate the indications for device implantation between primary (prevent sudden cardiac death in HF patients) vs. secondary prevention (prevent subsequent episodes in patients with ventricular arrhythmia), though secondary prevention indications (e.g., channelopathies) would be rare in the VA due to military service exclusions. Furthermore, BNP and NT-proBNP were not routinely measured, and the ordering itself may indicate HF decompensation, limiting our ability to make an apples-to-apples comparison with the trial population. Lastly, the magnitude of the intention-to-treat effect may vary substantially depending on the degree of treatment adherence. While 82.4% of participants randomized to the sacubitril/valsartan arm remained on the assigned treatment and completed the trial(49), only 52.1% of patients in our sample adhered to sacubitril/valsartan therapy throughout the follow-up. However, given the high comorbidity burden and lower treatment adherence in our sample, we expected the real-world effectiveness of sacubitril/valsartan to be attenuated relative to the efficacy observed in the trial. This also suggests that comparing the adherence-adjusted per-protocol effects between the trial and the observational emulation may be more appropriate for benchmarking.(50) Further, we did not compare across different ML algorithms. Instead, we focused on GBM, the most commonly used ML methods in the literature, to illustrate the paradoxical goal of predictive versus causal objectives, and the risk of bias when propensity models were misunderstood as predictive models.

In summary, ML-based propensity score estimation did not outperform logistic regression with the same pre-specified confounder set in this benchmark study. Residual confounding from omitted or partially adjusted variables likely contributed to this discrepancy. And when additional predictors were combined with automated feature selection, ML approach introduced bias. Therefore, while ML methods may add value with complex interactions, they are not a panacea. Ultimately, careful study design, reasoning, and subject-matter expertise—rather than algorithmic complexity and shortcuts—remain the foundation for producing reliable and causally interpretable real-world evidence from observational data.

## ACKNOWLEDGEMENTS

The views expressed in this manuscript are those of the authors and do not necessarily represent the views of the National Heart, Lung, and Blood Institute; the National Institutes of Health; the U.S. Department of Health and Human Services; or the U.S. Department of Veterans Affairs.

## STATEMENTS & DECLARATIONS

### Author Contributions

All authors contributed to the study conception and design. Material preparation, data collection and analysis were performed by Kaicheng Wang. The first draft of the manuscript was written by Kaicheng Wang and all authors commented on previous versions of the manuscript. All authors read and approved the final manuscript.

### Ethics Approval

This study was approved by the Research & Development Committee of the VA Connecticut Healthcare System and a waiver of informed consent was granted (IRBNet# 1582934).

### Data Availability

The data used for this study cannot be shared publicly due to privacy regulations, but are available with an approved study protocol by the U.S. Department of Veterans Affairs (https://www.virec.research.va.gov/). All SQL queries and analytical code are available at https://github.com/kw375/ML_PS.

