## Supplementary for "Machine Learning Versus Logistic Regression for Propensity Score Estimation: A Benchmark Trial Emulation Against the PARADIGM-HF Randomized Trial"

**Supplemental Table 1. Key Protocol Components of the Device-Stratified PARADIGM-HF Index Trial, Target Trial Specification and Emulation Using Observational Data from the U.S. Department of Veterans Affairs, 2016-2020.**

|  | Index Trial | Target Trial Specification | Target Trial Emulation |
| --- | --- | --- | --- |
| Eligibility criteria | - Implanted with an ICD or CRT-D - Age ≥ 18 years old - NYHA class II-IV at screening with a reduced LVEF ≤ 35% measured within the past 6 months - BNP ≥ 150 pg/ml (NT-proBNP ≥ 600 pg/ml) at Visit 1 OR BNP ≥ 100 pg/ml (NT-proBNP ≥ 400 pg/ml) and a HFH within the last 12 months - On stable dose of an ACEI or ARB equivalent to enalapril 10mg/d for at least 4 weeks before screening - On stable dose of a β-blocker for at least 4 weeks before screening unless contraindicated or not tolerated - Had no unacceptable side effects of the target doses of enalapril and sacubitril/valsartan during run-in period (6-8 wks) - No history of angioedema, heart transplant/LVAD within 3 months - No history of ACS, stroke, TIA, CABG, PCI or carotid angioplasty within the 3 months prior to screening - Not contraindicated or intolerance with ACEIs, ARBs or NEP inhibitor - Not currently decompensate, hypotension, impaired eGFR, hyperkalemia, or other significant conditions | - Implanted with an ICD or CRT-D* - Age ≥ 18 years old - Diagnosed with HF and a LVEF ≤ 35% - BNP ≥ 150 pg/mL or NT-proBNP ≥ 600 pg/mL - New or prevalent user of ACEI/ARB - Received GDMT (β-blocker, MRA or SGLT2i) - No run-in period - No history of LVAD or heart transplant - Not contraindicated or intolerance with ACEIs, ARBs or NEP inhibitor. - Not currently decompensate, hypotension, impaired eGFR, hyperkalemia, or other significant conditions - Implantation after January 1, 2016 - Study period between January 1, 2016 and September 30, 2020 | Same as index trial apart from:   - Assume no contraindications or not at high risk of adverse events of ACEI/ARB or sacubitril/valsartan |
| Treatment strategies | - Sacubitril/valsartan, 200mg/160mg bid - Enalapril, 10mg bid   Completely discontinued when consent withdrawal or detrimental to one’s well-being, otherwise be reintroduced after temporary discontinuation. | - Initiation/augmentation with sacubitril/valsartan and continuation over follow-up until contraindicated - Initiation/continuation of the ACEI/ARB had been receiving over follow-up until contraindicated | In addition to target trial:   - Discontinuation defined as a 30-day or longer gap between successive prescriptions - New user as not used an ACEI/ARB in the last 180 days |
| Treatment assignment | Afte run-in period (2 wks of enalapril and additionally 4-6 wks of sacubitril/valsartan), participants were randomly assigned in a 1:1 ratio | Participants were randomly assigned to a treatment and were aware of the strategy | Classified patients to strategy based on their compatible data at baseline:   - Sacubitril/valsartan: first prescription - ACEI/ARB: first fill in the study period |
| Outcome | - All-cause mortality - CV-specific morality - Heart failure hospitalization - KCCQ score - New onset atrial fibrillation - Renal dysfunction | All-cause mortality | Same as targe trial |
| Follow-up | After randomization, site visits every two to eight weeks in the first 4 months, then every four months, ends at death, withdrawal, lost-to-follow-up, when 2,410 patients have experienced a composite event or pre-specified early-stopping criteria for efficacy or safety are met (up to 51 months by March 31, 2014) | Starts at treatment assignment and ends at the date of death, 51 months after baseline, or administratively end of follow-up on September 30, 2020, whichever happened first. | Same as targe trial |
| Causal contrast | Intention-to-treat | Intention-to-treat | Same as target trial |
| Statistical analysis | Kaplan-Meier estimates and Cox proportional hazards models with treatment and region as fixed effect | RD and RR estimated from pooled logistic regression with IPTW for baseline covariates | Same as target trial |

Abbreviations: ACEI, angiotensin converting enzyme inhibitor; ACS, acute coronary syndrome; ARB, angiotensin II receptor blocker; BNP, brain natriuretic peptide; CABG, coronary artery bypass grafting; CRT-D, cardiac resynchronization therapy with defibrillator; CV, cardiovascular; eGFR, estimated glomerular filtration rate; ICD, implantable cardioverter defibrillator; KCCQ, Kansas City Cardiomyopathy Questionnaire; LVAD, left ventricular assist device; LVEF, left ventricular ejection fraction; MRA, mineralocorticoid receptor antagonist; NEP, neprilysin; NT-proBNP, N-terminal pro-brain natriuretic peptide; NYHA, New York Heart Association; PCI, percutaneous coronary intervention; SGLT2i, sodium-glucose cotransporter 2 inhibitors; TIA, transient ischemic attack.

**Supplemental Table 2. Variable Definitions**

|  | **Source** | **Definition** | **Form** | **Code** |
| --- | --- | --- | --- | --- |
| ***Exposure*** | | | | |
| Angiotensin converting enzyme inhibitor | CDW pharmacy | Benazepril, captopril, enalapril, fosinopril, lisinopril, moexipril, perindopril, quinapril, ramipril, trandolapril | Binary |  |
| Angiotensin II receptor blocker | CDW pharmacy | Azilsartan, candesartan, irbesartan, losartan, olmesartan, telmisartan, valsartan | Binary |  |
| Sacubitril/valsartan | CDW pharmacy | Sacubitril/valsartan | Binary |  |
| ***Outcomes*** | | | | |
| All-cause mortality | Death Ascertainment file | Date of death from the Master Person Index (MPI) and the Social Security Administration Death Master File (SSA DMF), and VA healthcare data. | Time-to-event |  |
| ***Baseline covariates*** | | | | |
| Month of baseline | CDW pharmacy | The month when initiating treatment, beginning on January 1, 2016. | Natural spline with 2 knots |  |
| Age | CDW patient | Date of baseline subtracted by date of birth. | Natural spline with 2 knots |  |
| Gender | CDW patient | Self-identified gender identity. | Categorical | Men, Women |
| Race | CDW patient | Most commonly self-reported race values. | Categorical | American Indian/Alaska Native, Asian, Black, Multi-race, Native Hawaiian/Pacific Islander, White, Unknown |
| Ethnicity | CDW patient | Self-reported ethnicity. | Categorical | Hispanics, non-Hispanics |
| Residence | CDW patient | Attributed to the geocoded patient location. | Categorical | Urban, non-urban (rural, highly rural, unknown) |
| Area deprivation index | CDW patient | Attributed to the FIPS code of patient residential location, 2015 version | Natural spline with 2 knots |  |
| Employment status | CDW patient | Self-reported employment status | Categorical | Not employed, full-time employed, part-time employed, retired |
| Cigarette smoking | CDW health factor | Health factors mapped to distinct smoking status and further refined to one-per-patient considering most recent smoking health factor in relation to past smoking health factors. | Categorical | Never, current, former, unknown |
| Body mass index | CDW vital status | 703 multiplicate the most recent weight in lbs. prior to baseline, then divided by median height across all measures | Natural spline with 2 knots |  |
| Left ventricular ejection fraction | VINCI NLP Output | Percentage of total blood volume pumped out the heart with each beat. Data extracted from echocardiogram, cardiac magnetic resonance and single-photon emission computed tomography reports, or from clinical notes | Natural spline with 2 knots |  |
| Systolic blood pressure | CDW vital status | Most recent measures prior to baseline. | Natural spline with 2 knots |  |
| Creatinine | CDW lab chemistry | Most recent serum/plasma creatinine prior to baseline | Continuous |  |
| Estimated glomerular filtration rate |  | Calculated from creatinine using the 2021 CKD-EPI equation | Natural spline with 2 knots |  |
| Potassium | CDW lab chemistry | Most recent serum/plasma potassium prior to baseline | Natural spline with 2 knots |  |
| BNP/NT-proBNP | CDW lab chemistry | Most recent BNP/NT-proBNP prior to baseline | Continuous |  |
| Albumin | CDW lab chemistry | Most recent serum/plasma prior to baseline | Continuous |  |
| Hemoglobin | CDW lab chemistry | Most recent hemoglobin from CBC panel prior to baseline (excluding serum/plasma hemoglobin level) | Continuous |  |
| Comorbidity | CDW outpatient and inpatient | ICD-9/10 codes recorded during at least two outpatient encounters, or from any hospitalization discharge diagnoses prior to the baseline | Binary | Yes/no |
| *Atrial fibrillation* |  | ICD9: 427.3  ICD10: I48 |  |  |
| *Cardiomyopathy* |  | ICD9: 425, 429.83  ICD10: I42, I43, I51.81 |  |  |
| *Chronic kidney disease* |  | ICD9: 403, 404, 582, 583, 585, 586, 588, V42.0, V45.1, V56  ICD10: I12, I13, N03, N05, N18, N19, N25, Z49, Z94.0, Z99.2 |  |  |
| *COPD* |  | ICD9: 491-492, 493.2, 496  ICD10: J41-J44 |  |  |
| *Diabetes* |  | ICD9: 250  ICD10: E10, E11, E13 |  |  |
| *History of heart transplant* |  | ICD9: V42.1  ICD10: Z94.1 |  |  |
| *Hypertension* |  | ICD9: 401-405, 437.2  ICD10: I10-I16, I67.4, N26.2 |  |  |
| *Depression* |  | ICD9: 296.2, 296.3, 311  ICD10: F32, F33 |  |  |
| *Left ventricular assist device status* |  | ICD9:  ICD10: |  |  |
| *Myocardial infraction* |  | ICD9: 410  ICD10: I21, I22 |  |  |
| *Peripheral artery disease* |  | ICD9: 437.3, 440, 441, 443.1-443.9, 447.1, 557, V43.4  ICD10: I70, I71, I73.1-I73.9, I77.1, I79.1, I79.8, K55.1, K55.8, K55.9, Z95.82, Z95.9 |  |  |
| *Valvular heart disease* |  | ICD9: 394-397, 424, V42.2, V43.3  ICD10: I05-I08, I09.1, I34-I39, Z95.2-Z95.4 |  |  |
| *Anemia* |  | ICD9: 280-285  ICD10: D5_, D60-D64 |  |  |
| *Malignancy* |  | ICD9: 140-172, 174-195, 199-208  ICD10: C00-C43, C45-C76, C80-C96 |  |  |
| *DVT/PE* |  | ICD9: 415, 451.1, 451.2  ICD10: I26, I80.1-I80.3 |  |  |
| *Dementia* |  | ICD9: 290, 331.0, 331.1, 331.82, 331.83  ICD10: F00-F03, G31.0, G31.83, G31.84 |  |  |
| *Influenzas* |  | ICD9: 487, 488  ICD10: J09-J11 |  |  |
| *Non-pathological fracture* |  | ICD9: 800-829  ICD10: S02, S12, S22, S32, S42, S52, S62, S72 |  |  |
| *Cerebral vascular conditions* |  | ICD9: 430, 431, 433._1, 434._1, 435, 436  ICD10: I60, I61, I63, G45 |  |  |
| Medications | CDW pharmacy | Medications released from outpatient pharmacy and were taking at the baseline. | Binary | Yes/no |
| *β-blockers* |  | Acebutolol, atenolol, bisoprolol, carvedilol, labetalol, metoprolol, nadolol, nebivolol, pindolol, propranolol, timolol, sotalol |  |  |
| *MRAs* |  | Eplerenone, spironolactone |  |  |
| *SGLT2i* |  | Bexagliflozin, canagliflozin, dapagliflozin, empagliflozin, ertugliflozin, sotagliflozin |  |  |
| *Diuretics* |  | Bumetanide, furosemide, torsemide; bendroflumethiazide, chlorthalidone, chlorothiazide, hydrochlorothiazide, hydroflumethiazide, indapamide, metolazone, polythiazide |  |  |
| *Digitalis* |  | Digoxin |  |  |
| Cardiac resynchronization therapy | Carelink data from manufacture | Implantable cardioverter defibrillators compatible to deliver cardiac resynchronization therapy |  |  |
| Influenza vaccine | CDW immunization | CVX code in 15, 16, 88, 111, 125-128, 135, 140, 141, 144, 149-151, 153, 155, 158, 160, 161, 166, 168, 171, 185, 186, 194, 197, 200-202, 205, 231, 320 in the past 12 months. | Binary | Yes/No |
| Heart failure hospitalization | CDW inpatient | Heart failure hospitalization occurred in the past 12 months prior to baseline | Categorical | None, any |
| Time since heart failure diagnosis |  | Time since the initial diagnosis of heart failure | Continuous |  |
| Time since implantation |  | Time since device implantation |  |  |

“_” is an SQL regular expression as an placeholder following. Abbreviations: ACEI, angiotensin converting enzyme inhibitor; ARB, angiotensin II receptor blocker; BNP, brain natriuretic peptide; CDW, Corporate Data Warehouse; COPD, chronic obstructive pulmonary disease; DVT/PE, deep vein thrombosis and pulmonary embolism; MRAs, mineralocorticoid receptor antagonists; NT-proBNP, N-terminal pro-brain natriuretic peptide; SGLT2i, sodium-glucose cotransporter 2 inhibitors.

**Supplemental Figure 1. Standardized Mean Differences Before Adjustment and After Applying Inverse Probability Weights Estimated Using Traditional Logistic Regression and Generalized Boosted Model Methods with the Same Pre-Specified Confounders.**

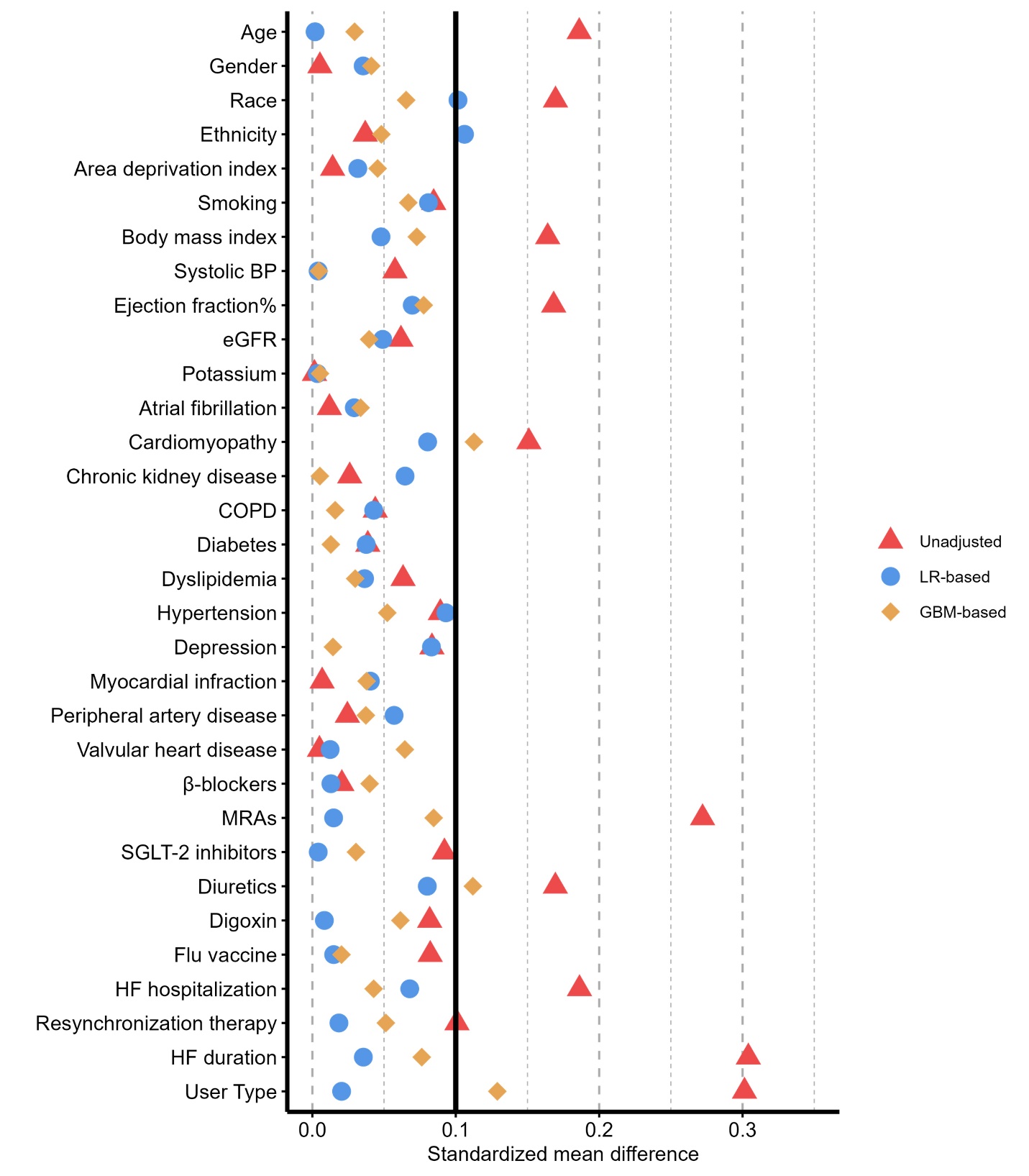

Abberetiations: BP, blood pressure; COPD, chronic obstructive pulmornary disease; eGFR, estimated glomerular filtration rate; GBM, generalized boosted model; HF, heart failure; LR, logistic regression; MRAs, mineralocorticoid receptor antagonists.

**Supplemental Figure 2. Variable Importance Plots for Covariates Used in the Generalized Boosted Model with (a) *Pre-Specified Confounders* and (b) Additional Predictors with Data-Driven Automated Selection.**

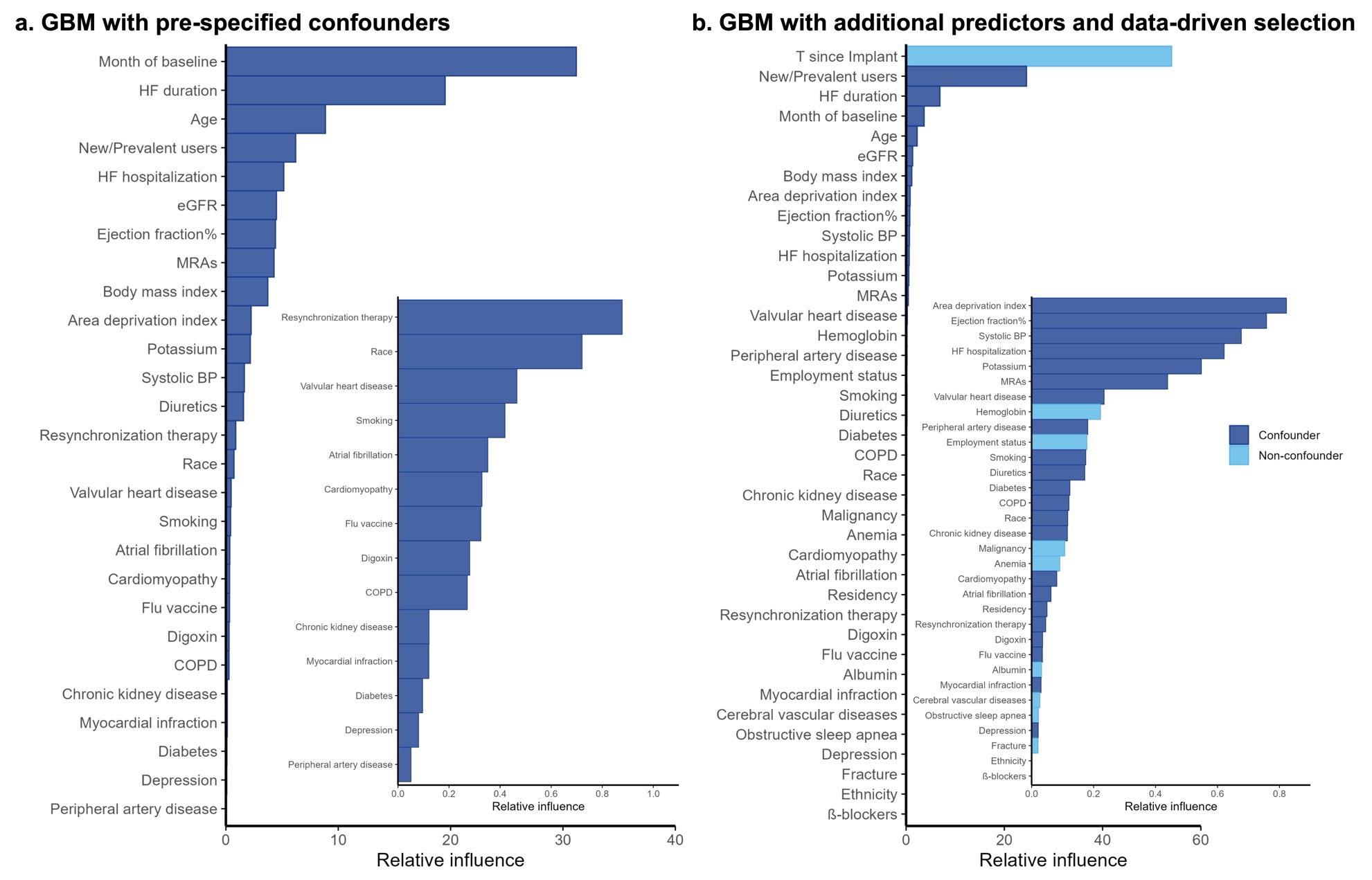

Abberetiations: BP, blood pressure; COPD, chronic obstructive pulmornary disease; eGFR, estimated glomerular filtration rate; GBM, generalized boosted model; HF, heart failure; LR, logistic regression; MRAs, mineralocorticoid receptor antagonists.
